# An Optimal Mass Transport Model for the Analysis of DCE-MRI and its Application to Breast Cancer Treatment Response

**DOI:** 10.1101/2024.11.05.24316768

**Authors:** Xinan Chen, Wei Huang, Amita Shukla-Dave, Ramesh Paudyal, Roberto Lo Gullo, Marcelina Perez, Katja Pinker, Joseph O. Deasy

**Affiliations:** Department of Medical Physics, Memorial Sloan Kettering Cancer Center, New York, USA; Advanced Imaging Research Center, Oregon Health & Science University, Oregon, USA; Department of Radiation Oncology, Corewell Health William Beaumont University Hospital, Michigan, USA; Department of Radiology, Memorial Sloan Kettering Cancer Center, New York, USA; Department of Radiology, Columbia University, New York, USA; Departments of Computer Science and Applied Mathematics & Statistics, Stony Brook University, New York, USA

**Keywords:** optimal transport, fluid dynamics, DCE-MRI, breast cancer

## Abstract

**Purpose:** Dynamic contrast-enhanced MR imaging (DCE-MRI) is widely deployed in cancer care and research, but the methods conventionally used to quantify contrast agent kinetics do not account the cross-voxel movement characterized by advection and diffusion. We hypothesized that unbalanced optimal mass transport could be used to quantify and visualize such contrast agent flows across tumor volumes.

**Methods:** We developed a computational fluid dynamics model termed the unbalanced regularized optimal mass transport (urOMT) model. We tested the urOMT on a multi-institutional dataset of 153 longitudinal DCE-MRI scans from 39 breast cancer patients treated with neoadjuvant chemotherapy (NACT.)

**Results:** The urOMT model can quantify dynamic fluid transport properties such as net speed, flux and rates of contrast entering and leaving the tumor (influx and efflux). The urOMT model can also visualize the trajectories and directions of net fluid flows. Quantitative metrics from urOMT exhibited distinct patterns that may be relevant to predicting pathological complete response (pCR) to NACT.

**Conclusion:** The urOMT model can be used to estimate and visualize local fluid flow in DCE-MRI breast cancer images. Model-based estimates of flux, influx and efflux should be tested as potential predictive imaging biomarkers to measure treatment effectiveness in patients treated with NACT. The urOMT model in principle has applicability to other cancer imaging use cases, but this will require further testing.

## 1 INTRODUCTION

Locally advanced breast cancer is often treated with neoadjuvant chemotherapy (NACT), aiming for the reduction of tumor size prior to definitive surgery. ^1,2^ However, patient response to NACT is highly variable. ^3,4^ Improved imaging methods that yield clinically useful prognostic information and effectively predict treatment response could potentially be used to adapt individualized treatment courses.

T1-weighted dynamic contrast-enhanced magnetic resonance imaging (DCE-MRI) has become a widely used imaging technique in many areas of cancer care and research, including breast cancer. ^5,6,7,8,9^ In DCE-MRI data acquisition, a chemically inert gadolinium-based contrast agent is usually injected intravenously into the patient. Before, during, and after the contrast administration, a series of MR images are acquired to identify the uptake and washout patterns exhibited by contrast agent for assessment of contrast kinetics. ^7,8^ Tumors generally demonstrate faster uptake and washout of the contrast agent compared to normal tissues, but in a spatially heterogenous pattern. ^10^

The term *advection* broadly refers to the net transport of directional bulk flows of a quantity, and can be mathematically characterized with a continuous velocity vector field with a local magnitude and direction. In the human body, advection is primarily driven by a gradient of pressure to enable the transport of fluids and gases essential for normal physiological functions. Distinct from advection, *diffusion* is directionless transport due to a gradient of concentration, and can be quantitatively described by the Fick’s law. During a DCE-MRI experiment, the contrast agent first arrives at the tumor within blood vessels, often entering increasingly smaller and disorganized capillaries in the tumor. Contrast agent extravasates blood vessels passively, a process called *influx*, thereupon entering the extravascular-extracellular tissue space. Contrast agent flow within the tissue space is non-advective and mainly via directionless diffusion. Eventually, the contrast agent passively exits the extravascular-extracellular space through a process called *efflux*, with some portion reabsorbed back to the blood vessels and the rest drained via lymphatic vessels. ^11,12,13,14^ Figure 1 shows a schematic of the aforementioned process. Advection mainly captures the net in-vessel bulk flows, which are microscopically varied, whereas diffusion captures the uncorrelated flow component in the interstitial space of the tissue.

**FIGURE 1.**
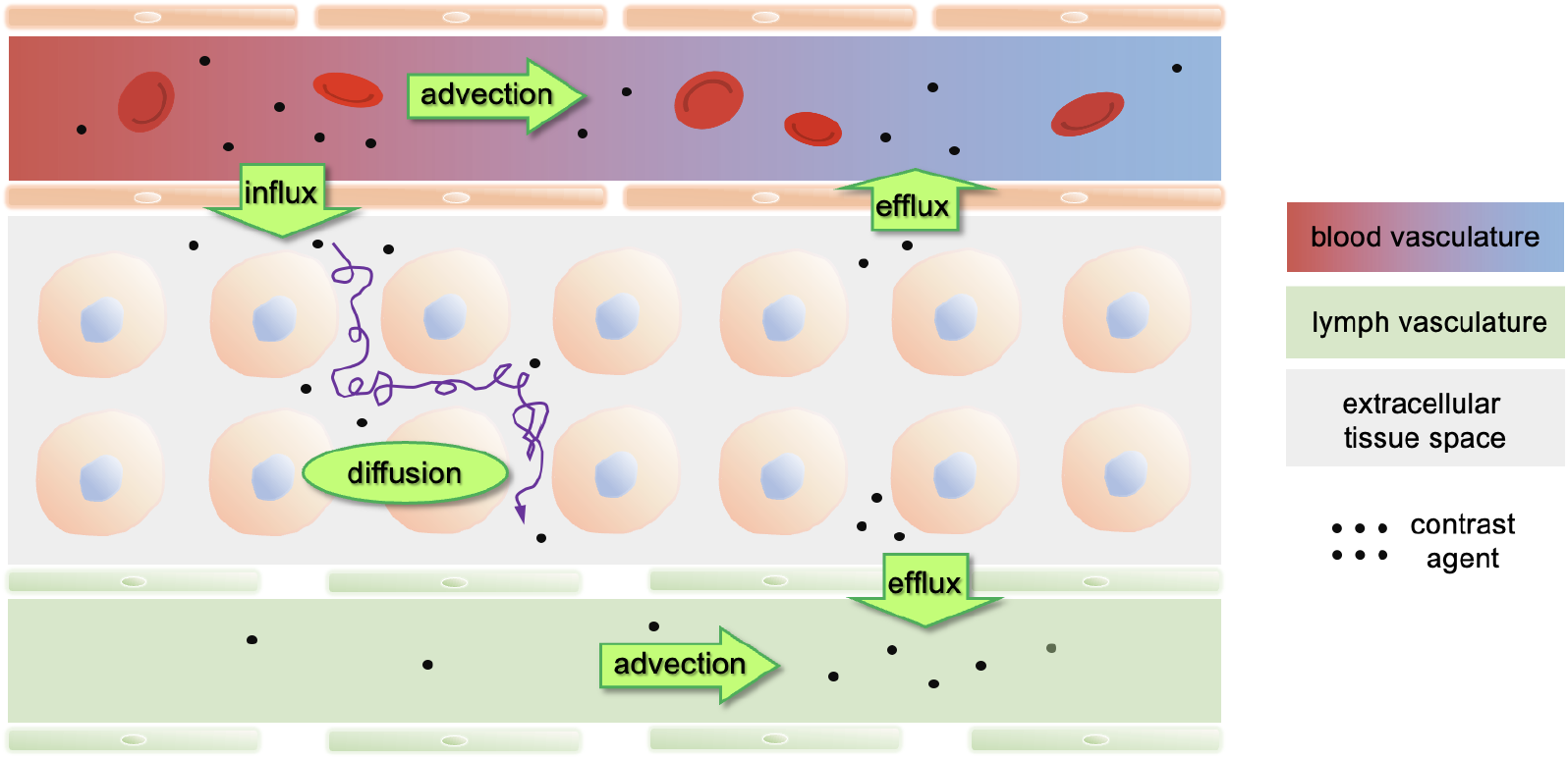
Schematic of the transport flows of the contrast agent after being intravenously administrated. Contrast agent molecules are delivered to the tumor via relatively large blood vessels which feed disorganized arterial microvasculature. Blood and contrast agent are passively transported into interstitial extracellular tissue regions in a process referred to as influx. Directionless diffusion of the contrast agent can occur in the extravascular-extracellular space, but is limited in range. The interstitial contrast agent is removed both through venous blood vessels or into the lymphatic drainage system (efflux). Individual contrast molecules may exit and enter the vascular system multiple times.

The DCE-MR images can be analyzed with semiquantitative (SQ) methods, which evaluate signal changes over time, ^15^ or quantitative methods, which perform pharmacokinetic (PK) modeling of tissue contrast concentration time-course data converted from signal time-course data. ^16,17,18,19^ The latter require high temporal resolution of the time-course data and quantification of the arterial input function. ^20^ Previous studies reported that the SQ and PK methods (e.g., extended Tofts model (ETM) and shutter speed model) exhibited potential for early prediction of breast cancer response to NACT by capturing the longitudinal changes of tumor contrast agent kinetics in response to NACT. ^21,22,23,24^

However, ETM ^25,26^ models only consider the direct exchange of contrast agent molecules between the vessels (considered one global reservoir) and the local interstitial tumor tissue in a given image voxel, but completely ignore any cross-voxel transport throughout the microvasculature and tissue characterized by advection and diffusion. Consequently, the transport of contrast agent within a tumor is not explicitly considered or modeled, and there is a great interest and unmet need to develop a new state-of-the-art model for the analysis of DCE data that includes the transport effects of advection and diffusion.

Our approach to address this problem is to apply optimal mass transport (OMT) theory to DCE-MRI data to track the cross-voxel flows in the tumor microenvironment. The OMT problem is concerned with finding the best transport plan from one initial mass distribution to another in the sense of minimized transportation cost. ^27,28,29^ In the formulation of Benamou and Brenier, ^29^ the OMT problem was, for the first time, framed in a fluid dynamics setting where the classic continuity equation is employed to describe the advective movement of fluid flows. A regularized version of the OMT model was later proposed ^30,31,32,33^ and has been widely applied in numerous research areas, including machine learning and deep learning ^34,35,36,37^ but also in molecular biology. ^38,39^ From the point of view of fluid dynamics, this regularization term can be equivalently treated as a diffusion term of a given quantity. In our previous work, we developed a regularized version of the optimal mass transport model (denoted rOMT) in computational fluid dynamics which employed an advection-diffusion equation to quantify and visualize glymphatic fluid behaviors in rat brains studied with DCE-MRI. ^40,41,42,43,44,45^ In order to further extend the validity of the model when contrast enters and leaves the imaged volume, we added a source term into the rOMT model to allow for unbalanced mass gain and loss, resulting in the unbalanced regularized OMT (urOMT) model. ^46^ The unbalanced version of OMT has been studied in other applications ^47,48,49^ including image processing ^50,51^, neuroscience ^46^, and cell biology. ^52,53^

As continuing work of [46, 54, 55], here we employed the urOMT model, of which the numerical method was developed and detailed in Chen et al., [46] to analyze breast cancer DCE-MRI data. In the urOMT model, fluid behavior is governed by a fluid dynamic partial differential equation:

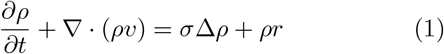

where *ρ* = *ρ*(*t, x*) is the concentration of a given quantity, *v* = *v*(*t, x*) is the velocity field of the quantity, *σ* is the constant diffusion coefficient of the quantity, and *r* = *r*(*t, x*) is called the source variable which quantifies the rate of mass gain (when *r >* 0) or loss (when *r <* 0) of the quantify. Optimal mass transport seeks to minimize an objective function while respecting the boundary conditions and the fluid dynamic equation. The urOMT algorithm solves for the optimal *v* and *r* which, under the context of cancer microenvironment measured by DCE-MRI, are the net velocity field and the rate of local influx or efflux rates of the injected contrast agent, respectively.

In this study, we applied the urOMT model to longitudinal DCE-MRI data sets obtained from breast cancer patients treated with NACT to quantify and visualize the fluid behaviors of the tumor microenvironment. The derived fluid dynamics metrics over the course of treatment were compared to outcomes of pathologic response as potential biomarkers of therapeutic response.

## 2 METHODS

### 2.1 DCE-MRI Data Acquisition and Pre-Processing

To investigate the applicability of urOMT to breast cancer treatment response, we analyzed a total of 153 longitudinal DCE-MRI data sets from 39 breast cancer patients treated with NACT. The data were obtained from a prospective single-center IRB-approved clinical study. ^56^ Most patients were consented to undergo four DCE-MRI scans: before NACT (baseline visit 1, denoted V1), after the first cycle of NACT (V2), at the mid-point of NACT (V3, usually after three or four NACT cycles) and after the completion of NACT but before surgery (V4). For DCE-MRI data acquisition, 28-32 frames of axial volumetric images with bilateral full breast coverage were acquired over a scan time lasting ∼ 10 minutes. See Ref. [56] for more details on DCE-MRI acquisition.

Standard of care pathological analysis of tumor specimens from surgery classifies NACT response outcome as either pathologic complete response (pCR, 8 patients) or non-pCR (31 patients). Here, a pCR is defined as the absence of residual tumor. Figure 2 displays the DCE-MRI images of one pCR patient and one non-pCR patient across four MRI visits.

**FIGURE 2.**
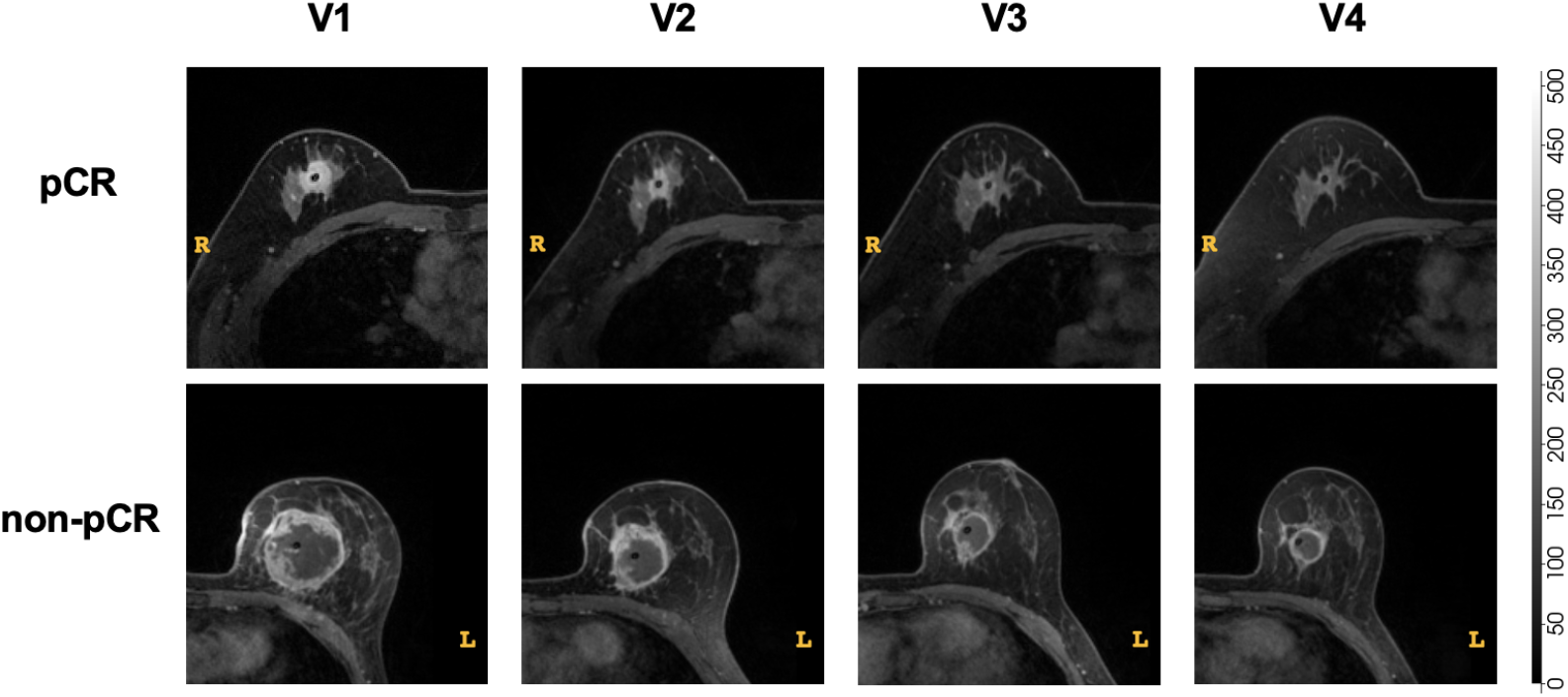
Cropped breast DCE-MRI image slice through the center of the tumor from a pCR patient (top row) and a non-pCR patient (bottom row) across 4 MRI visits, V1, V2, V3 and V4. The contrast-enhanced tumors are clearly visible for both patients at V1.

For each DCE-MRI study, the tumor contour (i.e., the tumor region of interest, ROI) was manually segmented out by fellowship trained dedicated breast radiologists with 1 and 9 years of experience on post-contrast images using 3D Slicer. ^57^

### 2.2 The urOMT Model

In this section, we describe the formal mathematical definition of the urOMT model and the corresponding numerical method developed for applications in 3D DCE-MRI images. ^40,42,46^

#### 2.2.1 Mathematical Formulation

The urOMT model used in this work can be formulated as follows. Given an initial mass density distribution *ρ*_0_(*x*) ≥ 0 and a final one *ρ*_1_(*x*) ≥ 0 defined on a bounded region Ω ⊆ ℝ ^3^, one seeks to solve the following optimization problem

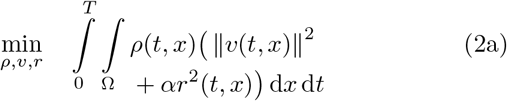

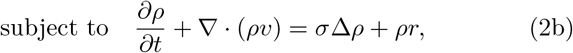

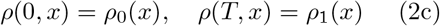

where *ρ*(*t, x*) : [0, *T*] × Ω → ℝ^+^ is the time-dependent mass density function; *v*(*t, x*) : [0, *T*] × Ω → ℝ^3^ is a net time-dependent velocity field which indicates the direction and magnitude of the advective transport; and *r*(*t, x*) is the time-dependent relative-source variable which controls the relative rate of instantaneous mass gain and loss (influx and efflux rates in the context of tumor microenvironment). The parameter *α >* 0 is the weighting parameter of the source term in the cost function (2a), and the parameter *σ >* 0 is the isotropic constant diffusion coefficient which allows the mass to be passively dispersed along its spatial gradient according to Fick’s law.

The equation (2b) is an unbalanced version of the usual advection-diffusion equation in fluid dynamics. In the language of transport phenomena, equation (2b) can be rewritten as

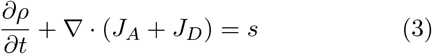

where *J*_*A*_ = *ρv* is the advective flux and *J*_*D*_ =− *σ* ∇ *ρ* is the diffusive flux describing the magnitude and direction of advective and diffusive transport, respectively; and *s* = *ρr* is the source describing the amount of entry (when *s >* 0) or the exit (when *s <* 0) of the given quantity into or out of the system per unit time. Note that *J*_*A*_ and *J*_*D*_ do not change the total amount of mass of the quantity in the entire process because advection and diffusion are both subject to the mass conservation law. Thus, we call ∇ · (*J*_*A*_ + *J*_*D*_) the *balanced* components of transport. On the contrary, *s* = *ρr* does contribute to the variation of the local and global mass (of the contrast agent in the tumor) and consequently we refer to it as the *unbalanced* component of the transport. The first term in the cost function (2a) is analogous to the total kinetic energy of the transport process within the tumor. In contrast, the second term is denoted the Fisher-Rao term and can be said to relate to the information geometry. ^58^ The urOMT problem (2) thus solves for the optimal transport strategy as measured by *ρ, v* and *r* which together transform *ρ*_0_ into *ρ*_1_ via the transport equation (2b), subject to a minimized cost function (2a).

In the context of the tumor microenvironment measured by DCE-MRI, the concentration of the injected contrast agent, calculated with known longitudinal relaxivity from the DCE-MRI signal intensity, is modeled by the mass density *ρ* in the urOMT formulation (2). The velocity field *v* delineates the advective behavior (i.e., the net microvascular and lymphatic flow) in the analyzed region. As noted above, diffusion is characterized with a constant coefficient. Advection and diffusion together account for the cross-voxel exchange of the contrast agent. The relative-source *r* represents the relative rate of influx when *r >* 0 and efflux when *r <* 0. In summary, the urOMT model incorporates four types of transport: entry (influx) and exit (efflux) into and out of the tumor tissue volume, bulk advective flow within vessels and lymphatics, and localized diffusive flow that is extravascular and extracellular.

#### 2.2.2 Numerical Method

We first converted the DCE-MRI signal intensity into the concentration of the injected contrast agent, using the following equations for spoiled gradient recalled echo sequence (with minimum TE) and a linear relationship between longitudinal relaxation rate (*R*_1_ = 1*/T*_1_) constant and contrast agent concentration under fast exchange limit approximation: ^59^

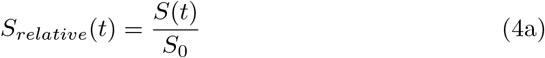

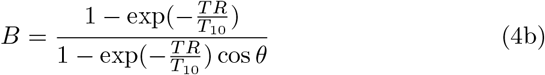

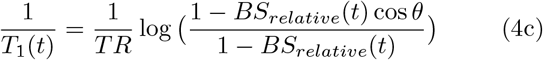

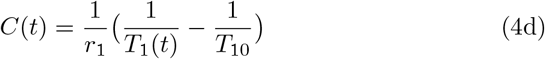

where *S*(*t*) is the MRI signal at time *t*; *S*_0_ is the baseline signal before administration of contrast agent; *TR* is the repetition time; T1 (t) is the longitudinal relaxation time constant at time t; *T*_10_ is the pre-contrast longitudinal relaxation time constant; *θ* is the flip angle; *r*_1_ is the longitudinal relaxivity of the contrast agent; *C*(*t*) is the concentration of contrast agent at time *t*.

*C*(*t*) is therefore represented by the mass density *ρ*(*t, x*) in the urOMT formulation (2). In order to deal with noise in the images, we first apply an edge-preserving filter to moderately smooth out image noise. In addition, in the numerical implementation, we solve the problem (2) with a free end point condition. Specifically, rather than a fixed end point condition in equations (2c) that *ρ*(*T, x*) = *ρ*_1_(*x*), we move this condition to the cost function to be minimized. The purpose is to avoid forcing the algorithm to match the noisy end-point exactly. Instead, this approach matches the end point image approximately. Consequently, the numerical formulation can be written as follows.

With two given 3D images, 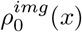 and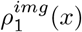, one solves

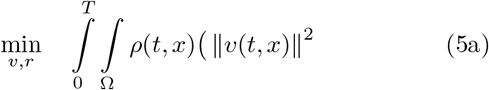

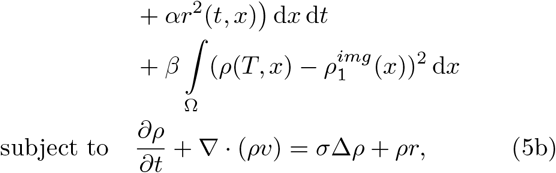

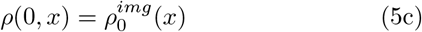

where *β >* 0 is the weighting parameter for the fitting term in the cost function.

Here we briefly outline the numerical solution scheme used. One can find more details in Ref. [46]. Note that a bold font is used to denote discretized flattened vectors. For the numerical discretization, the cubic region Ω is divided into a cell-centered grid of size *n*_1_ *×n*_2_*×n*_3_ with uniform length Δ*x*, Δ*y* and Δ*z*, and then *n* = *n*_1_*n*_2_*n*_3_ is the total number of voxels. The time interval [0, *T*] is discretized into *m* equal intervals of length Δ*t* = *T/m*. We set *t*_*i*_ = *i* Δ*t* for *i* = 0,, *m*. Set 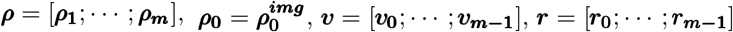 where each ***ρ***_*i*_ denotes the intensity at *t* = *t*_*i*_ and each pair of ***v***_*i*_ and ***r***_*i*_ denotes the velocity field and relative-source transforming ***ρ***_***i***_ to ***ρ***_***i*+1**_.

The discrete version of the problem (5) is therefore

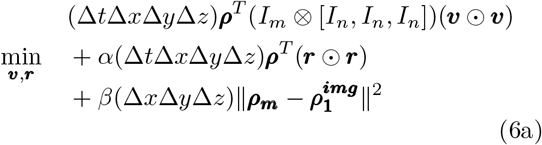

subject to ***ρ***_***i*+1**_ = *L*^−1^*S*(***v***_***i***_)*R*(***r***_***i***_)***ρ***_***i***_, for *i* = 0, …,*m* – 1

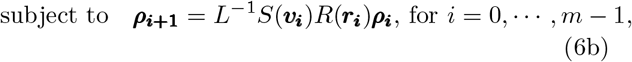

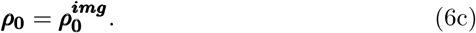

In equation (6a), ⊗ is the Kronecker tensor product; is the Hadamard product; *I*_*k*_ is the *k*-dimensional identity matrix; ||·|| is the *L*^2^ norm of a vector. Equation (6b) numerically solves the unbalanced advection-diffusion equation (5b) where an operator-splitting method is employed. Here, *S*(***v***_***i***_) is the averaging matrix in the advection step to redistribute mass according to ***v***_***i***_; *L* is the matrix for the diffusion operator; and *R*(***r***_***i***_) is the matrix for the mass gain and loss step.

##### Algorithm 1

Pseudocode for urOMT algorithm

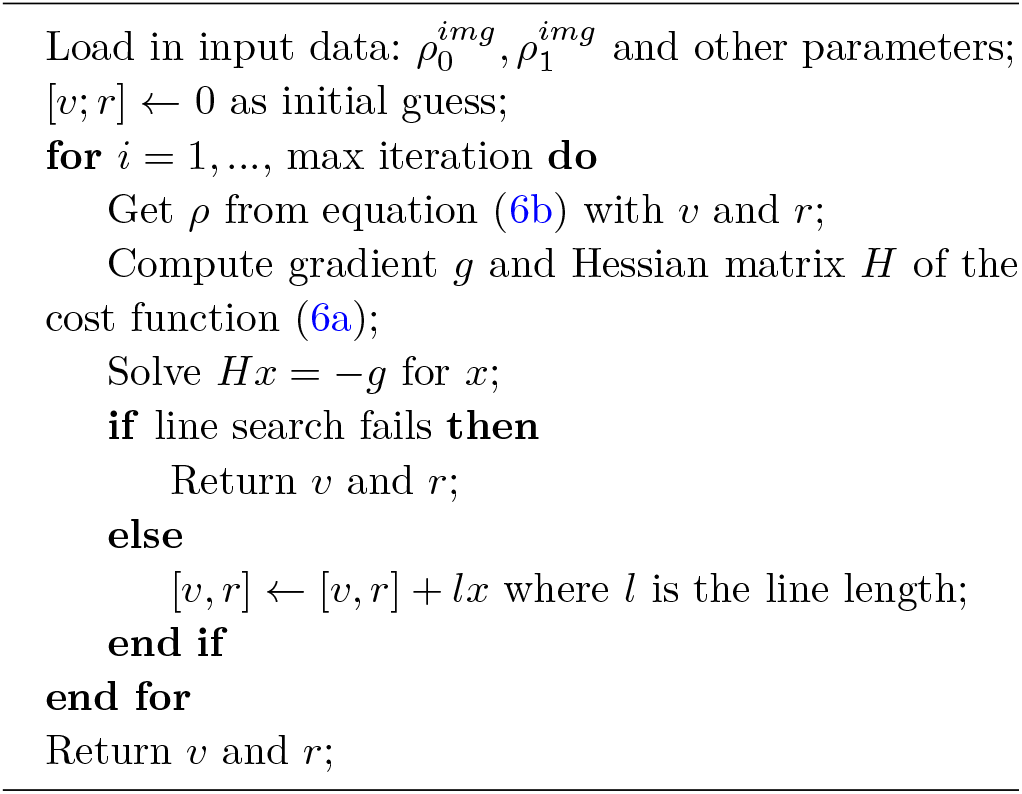

To numerically solve the discrete problem (6), we use a Gauss-Newton iterative algorithm. See the pseudocode in Algorithm 1. We run Algorithm 1 repeatedly between each pair of adjacent acquired DCE-MRI images to derive a series of prolonged velocity fields and relative-sources. Specifically, for each DCE-MRI study consisting of *q* images 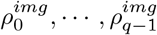, the urOMT algorithm gives the optimal velocity fields *v*_0,1_, …, *v*_*q*−2,*q*−1_ and relative-sources *r*_0,1_, …, *r*_*q*−2,*q*−1_, where *v*_*i*−1,*i*_ denotes the velocity and *r*_*i*−1,*i*_ denotes the relative-source transforming 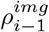 into 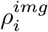 for *i* = 0, …, *q* − 2.

### 2.3 The Eulerian and Lagrangian Post-processing

With the model outputs, we then develop two post-processing methods, the Eulerian and the Lagrangian post-processing, to reveal and visualize the properties of the fluid behaviors in tumors. The difference between the two methods lies in the coordinate an observer chooses to observe the dynamic system.

The Eulerian method employs a fixed coordinate with which we can extract quantitative metrics as defined in Table 1 to reflect the local fluid properties. For example, Péclet is a dimensionless number to reflect the relative intensity of advection over diffusion. These metrics can be computed between each pair of adjacent image frames, and by connecting them together we have the corresponding time-varying metrics over all *q* −1 iterations.

**TABLE 1.** Quantitative metrics extracted from the Eulerian post-processing method.

| Name | Math Expression | Unit | Interpretation |
| --- | --- | --- | --- |
| speed | $\ v\ $ | mm/min | norm of the velocity field |
| flux | $\ \rho v\ $ | mm·mmol/(L · min)<sup>a</sup> | amount of mass (contrast agent) flowing per unit time, through a unit area; strength of advection |
| Péclet | $\frac{\ v\ }{\sigma \ \nabla \log(\rho)\ }$ | N/A | transport rate of advection/diffusion; the higher, the more advection-dominated; the lower, the more diffusion-dominated |
| influx | $\rho r$ where $r > 0$ | mmol/(L · min) | amount of mass (contrast agent) entering from the blood vessels to tissue per unit time; strength of influx |
| efflux | $\rho r$ where $r < 0$ | mmol/(L · min) | amount of mass (contrast agent) exiting from the tissue to the lymphatic and blood vessels per unit time; strength of efflux |

<sup>a</sup>mm is short for millimeter, mmol is short for millimole, and L stands for liter.

In contrast, the Lagrangian method employs a floating coordinate which enables us to track the trajectories of the cross-voxel flows contributed by net advection and diffusion from a seed point over time, which we name as the *pathlines*. Transport properties such as speed can also be computed along the pathlines to derive what we call the *speedlines*. By connecting the starting and terminal points of the pathlines, we derive the *displacement field* and these vectors may be used to visualize the direction and the distance a seed point has travelled in a neat manner. The metrics extracted from the Lagrangian post-processing method are summerized in Table 2.

**TABLE 2.** Metrics from tracking cross-voxel transport using the Lagrangian post-processing method.

| Metric Name | Interpretation |
| --- | --- |
| pathlines | binary trajectories/pathways of contrast agents |
| speed-lines | pathlines endowed with speed $\ v\ $ |
| displacement field | movement of cross-voxel transport over time; connecting the start and end points of pathlines |

More details can be found in Ref. [41, 46] about the post-processing methods.

### 2.4 Implementation Pipeline

Here we describe implementation of the entire analysis process in this study.

For each DCE-MRI study, we first manually segmented the tumor region and dilated the tumor region by 2 voxels (about 2 mm *×* 2 mm *×* 2.8 mm) to include some surrounding tissues as the region of interest (Figure 3 a). Instead of processing all MRI images in the DCE-MRI experiment, we started at the 5th image frame (approximately 30 s after contrast injection) and proceeded to the end of DCE-MRI acquisition skipping every other image frames, i.e., 5th → 7th →…, resulting in 12 ∼ 14 temporal 3D images to be analyzed. Starting with the 5th image provides a clear and stable signal. We then converted MRI signal intensity image within the ROI into contrast concentration image following equations (4a) - (4d) (Figure 3 b). Next, a 3D affine denoising filter ^60^ which preserves edges was used to smooth the concentration images within the ROI (Figure 3 c). The resulted smoothed series of volumetric data was processed by the urOMT algorithm ^61^ with *σ* = 0.002 (in numerical grid), *β* = 1000 and *α* = 30000 to obtain model outputs (Figure 3 d). Two post-processing methods were utilized to (i) generate temporal quantitative metrics detailed in Table 1 and (ii) track the trajectories of the cross-voxel contrast agent movement over the analyzed time window whose metrics are detailed in Table 2 (Figure 3 e).

**FIGURE 3.**
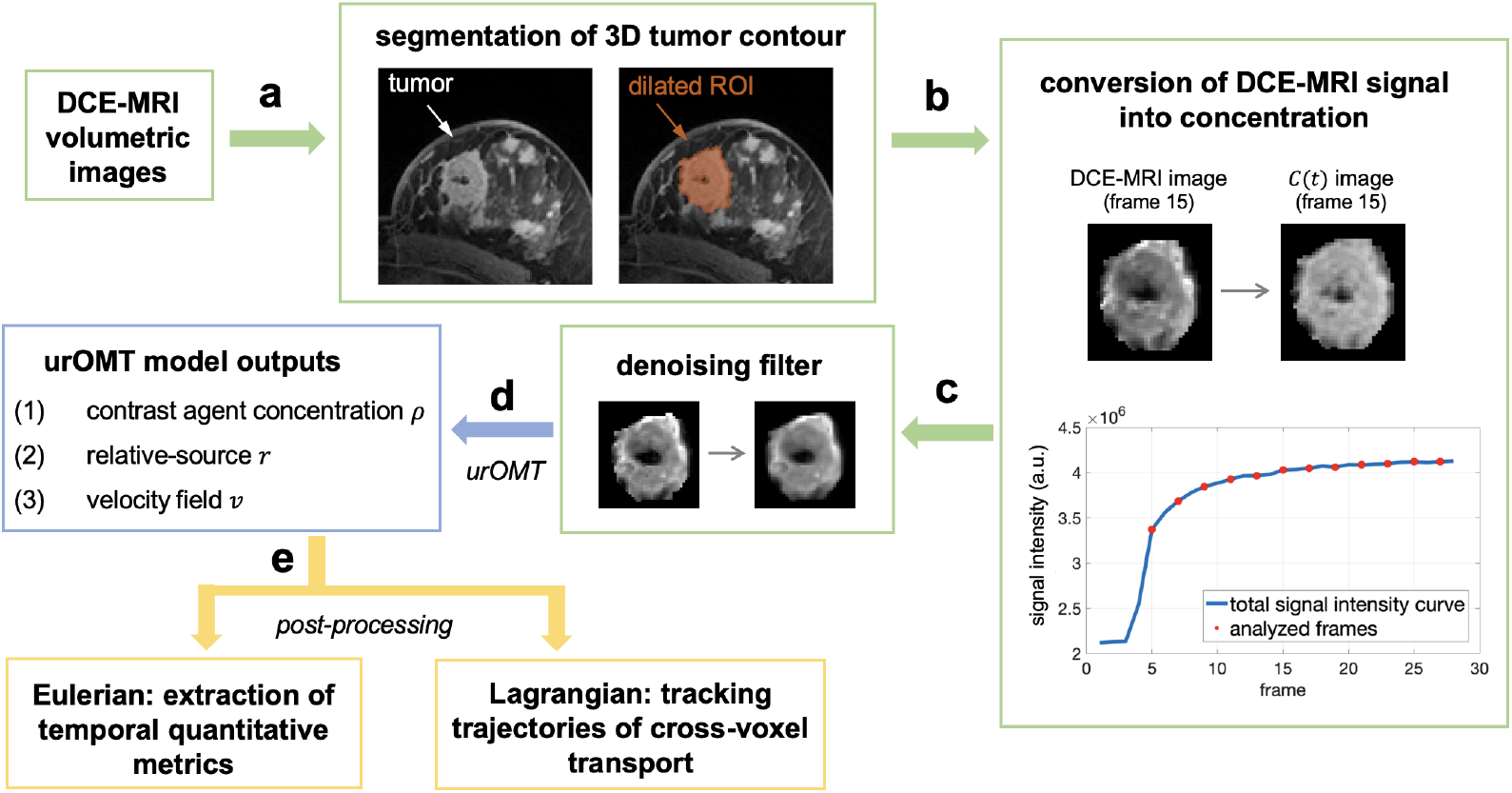
The urOMT analysis pipeline. The green represents pre-processing; blue represents the main algorithm; yellow represents post-processing. **a**. DCE-MRI images were first segmented by radiologists to derive the tumor contour which was further dilated by about 2 mm as the region of interest (ROI). **b**. DCE-MRI signal images within the ROI were converted into concentration *C*(*t*) images of contrast agent. **c**. The images within the ROI were further smoothed with a denoising filter to remove image noise. **d**. Pre-processed concentration images were then used as inputs for the urOMT model. **e**. Model outputs were post-processed to extract the temporal metrics listed in Table 1 and to track the trajectories of cross-voxel transport whose metrics are listed in Table 2.

### 2.5 Statistical Analysis

In the Eulerian post-processing, we used two-sample *t*-test assuming unequal variances to evaluate the difference between pCR and non-pCR patients.

## 3 RESULTS

### 3.1 urOMT Generates Dynamic Transport Metrics and Shows the Directional Trend of Fluid Flows

For illustration, we show detailed results for one representative non-pCR breast cancer patient at the baseline (V1) in Figure 4. The dynamic metrics illustrate the changes of the tumor fluid properties over the analyzed time window which lasted about 7 minutes (Figure 4 b). The times shown in Figure 4 b indicate the approximate number of seconds from the start of DCE-MRI acquisition. For this particular study, contrast agent injection at 2 mL/s was initiated approximately 80 s after the start of DCE-MRI acquisition. Speed was very high in the first two to three DCE frames that were analyzed by the urOMT algorithm, which was rapidly decreased in the later frames. For the flux and Péclet metrics, it is clear that the flows in the tumor were stronger and more advection-dominated near the ring-shaped boundary than the central region of the tumor. Influx was intense at first (colored dark red), indicating that the contrast agent was actively entering the tumor tissue. In later frames, efflux increases (colored light blue), indicating a slow drainage of the contrast agent from the tumor tissue to the lymphatics and venous blood flow. The Lagrangian results display cross-voxel trajectories of the contrast agent. In Figure 4 c-d, the pathlines and its corresponding displacement field indicate that the contrast agent flow primarily began as influx in a ring-shaped bordered region and mostly moved inwards to the center of the tumor. From the speed-lines, we can observe that speed of the flow was higher in the boundary than in the center of the tumor (Figure 4 e). From these results, we infer that in this case the tumor boundary was much more well-vascularized and that the center of the tumor was more diffusion-dominated and less active, indicating poor microvascularization or necrosis.

**FIGURE 4.**
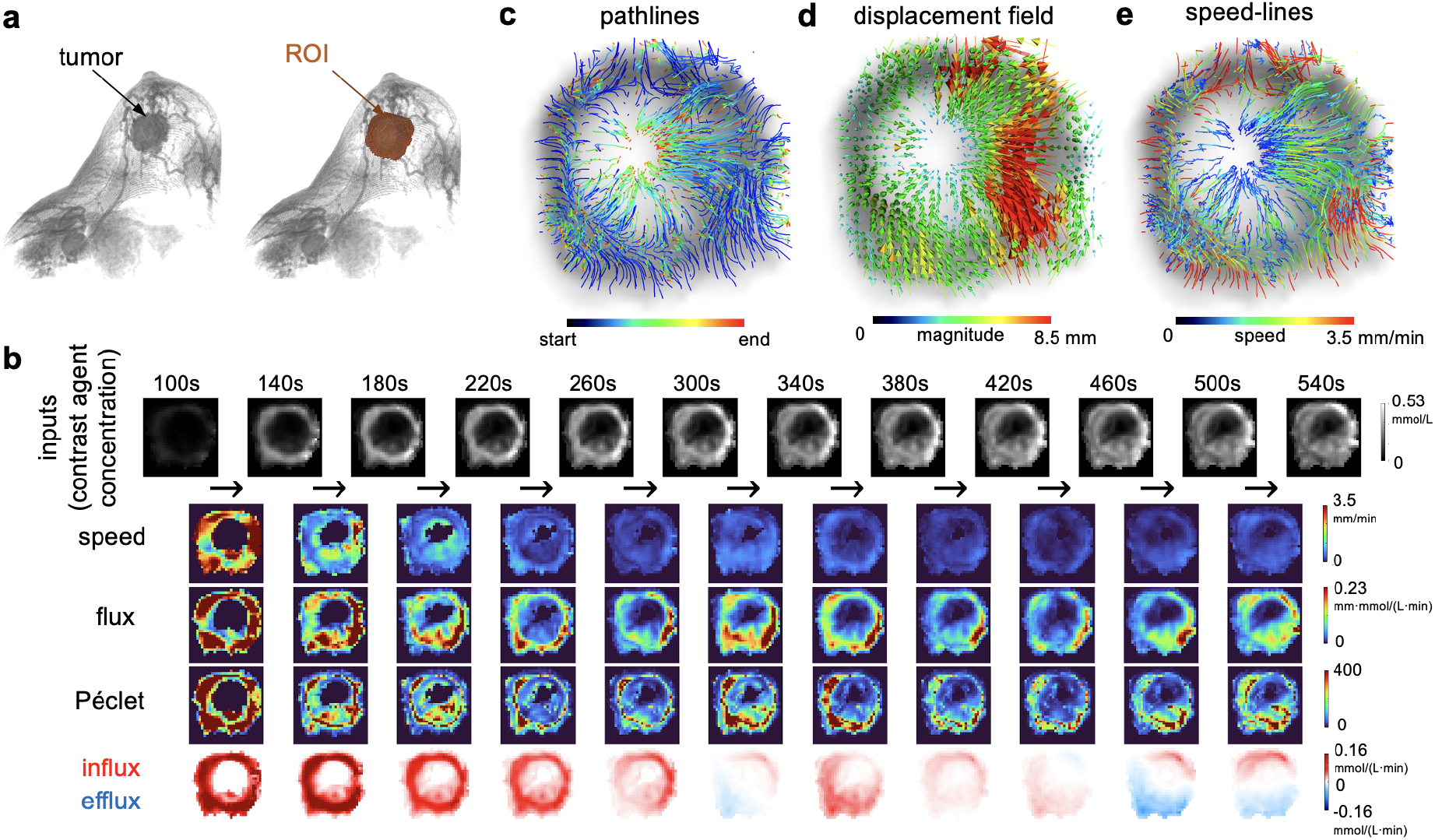
Results from a representative non-pCR patient at baseline (V1). **a**. 3D rendered post-contrast MRI image with the ROI of the tumor region color-coded in orange. **b**. Input contrast concentration images in the tumor ROI (first row) and dynamic results of quantitative metrics from the Eulerian post-processing (bottom four rows). The times shown in the first row are approximate number of seconds after the start of DCE-MRI acquisition. The images are shown at the same centered slice in 2D. **c-e**. Tracked cross-voxel flows over the analyzed time window from Lagrangian post-processing. The results are overlaid on a gray-scale concentration image and are shown in projection from the middle 5 slices of the tumor.

In Figure 5, the Lagrangian results of another non-pCR patient at baseline (V1) are shown. In some locations the fluid flow converged and at other locations diverged. Moreover, the speed was also unevenly distributed inside of the tumor. It seems likely that the tumor vasculature was robust in some regions (the yellow solid boxes) but mostly inactive in others (white dashed boxes) (Figure 5 b). A 3D-rendering video of the results can be found at Github.

**FIGURE 5.**
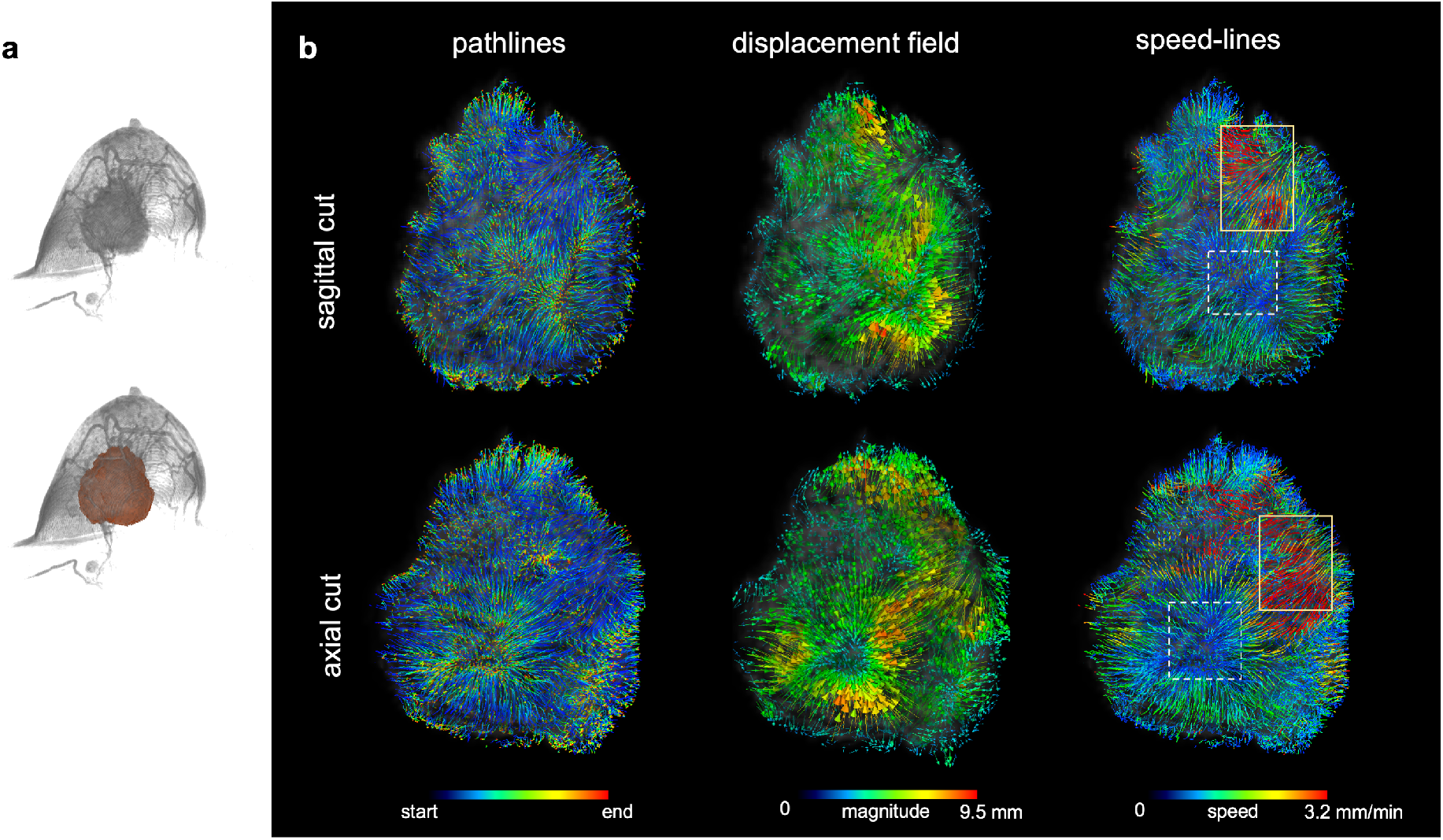
Heterogeneity of fluid flows can be visualized by the Lagrangian post-processing method. **a**. 3D rendered post-contrast MRI image of the tumor from a non-pCR patient at baseline (V1) with the ROI region color-coded in orange. **b**. Corresponding 3D rendered Lagrangian results (pathlines, displacement field and speed-lines) when the tumor is cross-sectioned by the central sagittal plane (top row) and the central axial plane (bottom row). The yellow solid boxes are where the tumor flow was the most active and the white dashed boxes are where the tumor flow was the most inactive.

### 3.2 urOMT-derived metrics as potential biomarkers of therapeutic response

Next, we computed the temporal average of flux, influx and efflux for all MRI visits. We observed that for the pCR patients the average flux, influx and efflux were all decreased compared to the baseline (V1), presumably due to the impact of NACT on the microvasculature. In contrast, the non-pCR patients seemed to be more likely to experience an increase in some metrics during NACT (at V2, V3 and V4). Figure 6 shows longitudinal changes of the time-averaged metrics for one pCR and one non-pCR patient. For the pCR patient, in addition to the shrinkage of the tumor, all the metric values decreased at the three visits (V2, V3 and V4) after NACT initiation compared to baseline (V1). However, for the non-pCR patient, there was a substantial elevation of flux and efflux at V3 and V4, although at V2 the metrics were slightly decreased compared to baseline. This suggests possible tumor resistance to NACT after a moderate initial response to the first NACT cycle.

**FIGURE 6.**
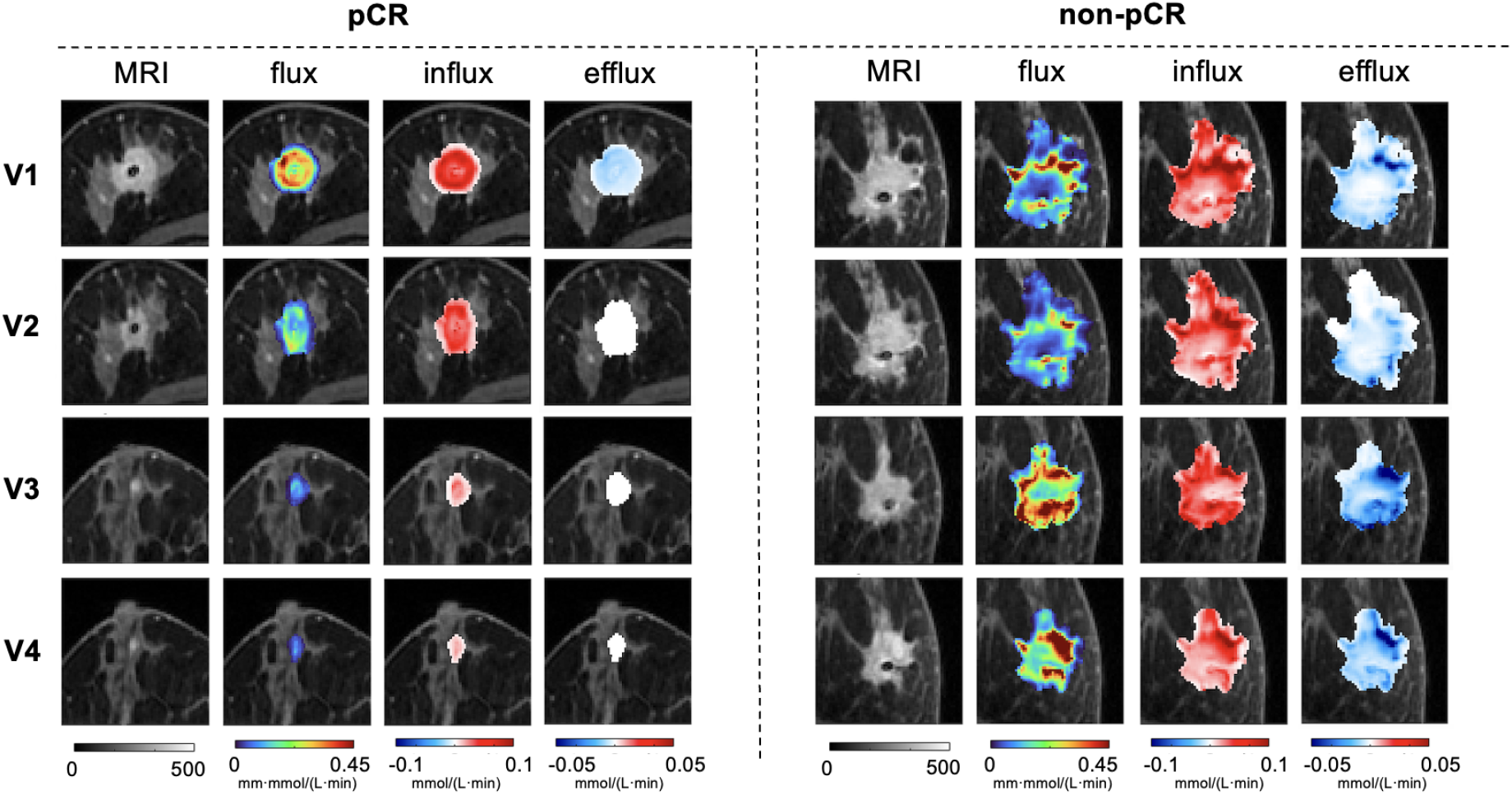
Comparison of temporal averages of quantitative flow metrics of one pCR patient (left) and one non-pCR patient (right) at four DCE-MRI visits before, during, and after NACT treatment. The first column is the last frame DCE-MRI images, while the other three columns are color parametric maps of the urOMT-derived metrics overlaid on the MR images.

For all 39 patients, we calculated the temporal and spatial averages of flux, influx and efflux for all 153 DCE-MRI scans and computed their percentage change from baseline (V1). To be specific, for V1→ V*i*, percentage change from baseline of a metric *M* is

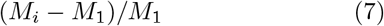

where *M*_*i*_ is the metric at V*i*. In this dataset, there are 3 patients whose V3 data are missing. These patients’ data were still included in the analysis for completeness, and we used the interpolated metric values at V2 and V4 as their estimated V3 metric values. There is one patient whose V3 and V4 were identified by radiologists as complete imaging response, thus no ROI was provided, for which we set their metrics to zero at V3 and V4. In Figure 7, we longitudinally plotted the percentage changes from baseline of average flux, influx, and efflux for all 39 patients. Note that flux at V3 was decreased to larger extent for pCR (−47.26%) than for non-pCR patients (2.28%, *p* = 0.0042). Flux was also lower at V4 in pCR (−51.70%) vs. non-pCR patients (−8.59%, *p* = 0.0111). Influx was reduced at V4 in pCR (−56.32%) than in non-pCR (−21.56%, *p* = 0.0473). Lastly, efflux was decreased more at V4 in pCR (−79.07%) than in non-pCR (−2.58%, *p* = 0.0189). Table 3 summarizes the the longitudinal percent change of the metrics values.

**TABLE 3.** Percent change from baseline of urOMT-derived metrics across all 39 patients at V2, V3 and V3.

| | | pCR ($N = 8$) | | non-pCR ($N = 31$) | | |
| --- | --- | --- | --- | --- | --- | --- |
| Metrics | Visit | mean | STD | mean | STD | $p$-value |
| flux | V1 → V2 | -0.2140 | 0.1762 | 0.0243 | 0.5624 | 0.0523 |
| | V1 → V3 | -0.4726 | 0.3049 | 0.0228 | 0.6341 | **0.0042\*\*** |
| | V1 → V4 | -0.5170 | 0.3251 | -0.0859 | 0.5680 | **0.0111\*** |
| influx | V1 → V2 | -0.1842 | 0.3395 | 0.0822 | 0.7144 | 0.1422 |
| | V1 → V3 | -0.4585 | 0.3437 | -0.1452 | 0.5736 | 0.0645 |
| | V1 → V4 | -0.5632 | 0.3433 | -0.2156 | 0.6200 | **0.0473\*** |
| efflux | V1 → V2 | -0.2648 | 0.7539 | 0.5034 | 1.8672 | 0.0832 |
| | V1 → V3 | -0.6038 | 0.5328 | -0.1086 | 1.0626 | 0.0777 |
| | V1 → V4 | -0.7907 | 0.3666 | -0.0258 | 1.5778 | **0.0189\*** |

\* $p < 0.05$ , \*\* $p < 0.005$ . A two-sample $t$ -test assuming unequal variances was used to assess difference between the pCR and non-pCR group.

**FIGURE 7.**
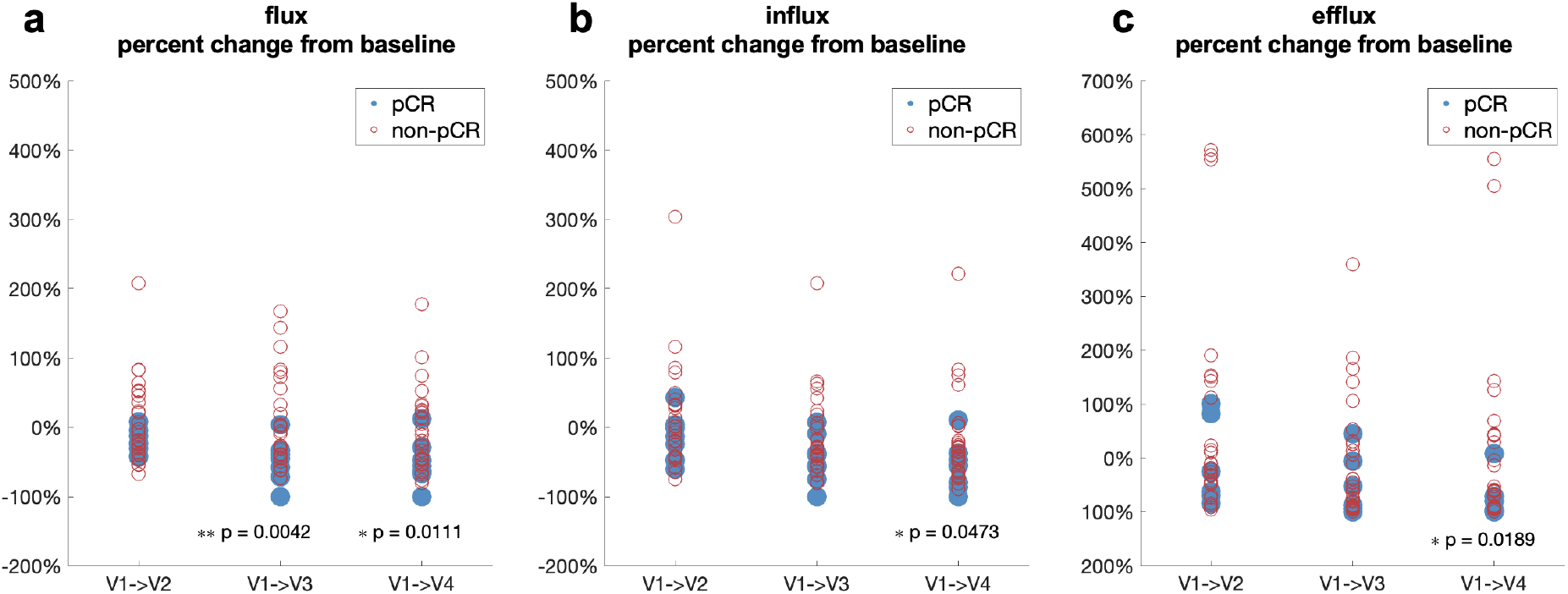
Percentage changes from baseline (V1) of average flux (**a**), average influx (**b**) and average efflux (**c**) for all 39 patients at V2, V3, and V4. Detailed statistics are listed in Table 3. A two-sample *t*-test assuming unequal variances was used to assess difference between the pCR and non-pCR group. ^*^*p <* 0.05, ^**^*p <* 0.005.

These observations suggested that although NACT reduced microvascular fluid transport in the breast tumors, the timing and extent of the effect were different between the pCRs and non-pCRs. If we simply identify patients whose average flux, influx and efflux were all decreased from V1 to all three follow-up visits as *urOMT-response-positive* and pCR as pathology test positive, we have the following confusion matrix comparing predictions by urOMT with pathologic response outcomes:

|  |  | Pathology Test |  | Total |
| --- | --- | --- | --- | --- |
|  |  | Pos. | Neg. |  |
| Prediction | Pos. | 6 | 7 | 13 |
|  | Neg. | 2 | 24 | 26 |
| Total |  | 8 | 31 | 39 |

By this definition, the accuracy of this simple response classifier was 77%. And the corresponding sensitivity and specificity are 75% and 77%, respectively.

## 4 DISCUSSION

Commonly used quantitative DCE-MRI data analysis approaches, for example, the Tofts-type methods ^25,26^ only allow for contrast influx and efflux, measured by biomarkers *K*^*trans*^ and *k*_*ep*_ to reflect volume transfer rate constants of the contrast agent for the passive transport mechanisms from the local arterial supply to tissue interstitial space and back into the draining venous vessels, respectively. However, their lack of considering cross-voxel flows characterized by advection and diffusion may lead to bias of the modeled results. For instance, Fluckiger et al. studied the effects of contrast molecule diffusion on the analysis of DCE-MRI data in realistic tissue domains and suggested incorporating a correction for the slow diffusion of the contrast agent. ^62^ In a recent study, Sinno et al. developed a modified Tofts-based approach for DCE-MRI analysis that explicitly models cross-voxel flows and demonstrated that ETM underestimates the effects of cross-voxel exchange. ^63^

In contrast, in addition to quantifying temporally and spatially resolved influx and efflux, the urOMT model can capture net directional bulk flows within the tumor volume. This is achieved via smooth continuous cross-voxel transport modeled by a combination of advection and diffusion. urOMT thus incorporates four transport motions under two categories, i.e., tumor blood supply (influx and efflux) and cross-voxel transport (advection and diffusion), to provide a more complete blood transport framework. Furthermore, the urOMT model enforces a continuity in space and time. Specifically, it can characterize the transport system by returning time-dependent dynamic metrics rather than time-independent constant metrics such as *K*^*trans*^ of Tofts-type models. This captures the changing behavior of contrast agent flow as it moves from an arterial supply phase to a venous/lymphatic removal phase. Finally, the urOMT model can track and visualize net trajectories of cross-voxel contrast transport over the analyzed time window, thus quantifying directional information on the fluid transport. To the best of our knowledge, this is the first model that traces detailed contrast flow patterns within a tumor.

In this preliminary study, we applied a mathematical model in computational fluid dynamics, called the urOMT model, to analyze longitudinal DCE-MRI data from breast cancer patients treated with NACT. As demonstrated by the initial results from 153 DCE-MRI data sets from 39 breast cancer patients, the urOMT-derived metrics have the potential to serve as imaging biomarkers to quantify and predict therapeutic responses. The applicability of urOMT is not limited to breast cancer, and could be applied to a wide variety of DCE-MRI studies of various cancer types.

The current urOMT model, however, is not without limitations. In particular, the model currently assumes that diffusion is constant across the tumor. This is certainly not strictly true, although in many cases diffusion may be a minor component of transport. This constraint could be relaxed in future versions. Besides, establishing the urOMT-derived metrics as robust biomarkers for cancer therapy response will require larger datasets for validation.

## Data Availability

All data produced in the present study are available upon reasonable request to the authors.

https://www.cancerimagingarchive.net/collection/qin-breast-dce-mri/

https://github.com/xinan-nancy-chen/urOMT

## ACKNOWLEDGMENTS

This study was supported by the NIH Cancer Center Support grant to MSK (P30 CA008748), NIH grant U01 CA154602, R01 CA248192, The Enid A. Haupt Endowed Chair Fund for Medical Physics, The Simons Foundation, and a Breast Cancer Research Foundation grant (BCRF-17-193).

We gratefully acknowledge Dr. Allen Robert Tannenbaum, whose mentorship and contributions have profoundly shaped this research. Dr. Tannenbaum sadly passed away before the completion of this work.

## 4.1 Author contributions

The project was conceived by X. C., W. H., and J. O. D. The theory, numerical method, code development of urOMT were done by X. C. The DCE-MRI data collection and processing were performed by W. H., A. S. D., and R. P. Tumor segmentation was performed by R. L. G., M. P. and K. P. The data interpretation was contributed by A. S. D., R. P., K. P. and J. O. D. The first draft of the manuscript was written by X. C. and all authors commented on previous versions of the manuscript. All authors read and approved the final manuscript.

## DATA AVAILABILITY STATEMENT

The code for the urOMT method can be found at Github (https://github.com/xinan-nancy-chen/urOMT) or at Zenodo. ^61^ The DCE-MRI data is taken from the study published by [56]. Data availability is limited due to privacy concerns and consent form limitations.

## REFERENCES

1. Charfare H, Limongelli S, Purushotham AD. Neoadjuvant chemotherapy in breast cancer. Journal of British Surgery. 2005;92(1):14–23.

2. Asaoka Mariko, Gandhi Shipra, Ishikawa Takashi, Takabe Kazuaki. Neoadjuvant chemotherapy for breast cancer: past, present, and future. Breast cancer: basic and clinical research. 2020;14:1178223420980377.

3. Antonini Marcelo, Mattar André, Richter Fernanda Grace Bauk, et al. Real-world evidence of neoadjuvant chemotherapy for breast cancer treatment in a Brazilian multicenter cohort: Correlation of pathological complete response with overall survival. The Breast. 2023;72:103577.

4. An Selena J, Duchesneau Emilie D, Strassle Paula D, et al. Pathologic complete response and survival after neoadjuvant chemotherapy in cT1-T2/N0 HER2+ breast cancer. NPJ Breast Cancer. 2022;8(1):65.

5. Mann Ritse M, Kuhl Christiane K, Moy Linda. Contrast-enhanced MRI for breast cancer screening. Journal of Magnetic Resonance Imaging. 2019;50(2):377–390.

6. Leithner D, Wengert GJ, Helbich TH, et al. Clinical role of breast MRI now and going forward. Clinical radiology. 2018;73(8):700–714.

7. Türkbey Bariş, Thomasson David, Pang Yuxi, Bernardo Marcelino, Choyke Peter L. The role of dynamic contrast-enhanced MRI in cancer diagnosis and treatment. Diagnostic and interventional radiology (Ankara, Turkey). 2010;16(3):186.

8. Udayakumar Durga, Madhuranthakam Ananth J, Doğan Başak E. Magnetic Resonance Perfusion Imaging for Breast Cancer. Magnetic Resonance Imaging Clinics. 2024;32(1):135–150.

9. Lee Jeongmin, Kim Sung Hun, Kang Bong Joo. Pre-treatment prediction of pathologic complete response to neoadjuvant chemotherapy in breast cancer: Perfusion metrics of dynamic contrast enhanced MRI. Scientific Reports. 2018;8(1):9490.

10. Choyke Peter L, Dwyer Andrew J, Knopp Michael V. Functional tumor imaging with dynamic contrast-enhanced magnetic resonance imaging. Journal of Magnetic Resonance Imaging: An Official Journal of the International Society for Magnetic Resonance in Medicine. 2003;17(5):509–520.

11. Schaaf Marco B, Garg Abhishek D, Agostinis Patrizia. Defining the role of the tumor vasculature in antitumor immunity and immunotherapy. Cell death & disease. 2018;9(2):115.

12. Wagner Marek, Wiig Helge. Tumor interstitial fluid formation, characterization, and clinical implications. Frontiers in oncology. 2015;5:115.

13. Bekisz Sophie, Baudin Louis, Buntinx Florence, Nöel Agnès, Geris Liesbet. In vitro, in vivo, and in silico models of lymphangiogenesis in solid malignancies. Cancers. 2022;14(6):1525.

14. Stewart Randolph H. A modern view of the interstitial space in health and disease. Frontiers in Veterinary Science. 2020;7:609583.

15. Jackson Alan, Li Ka-Loh, Zhu Xiaoping. Semi-quantitative parameter analysis of DCE-MRI revisited: Monte-Carlo simulation, clinical comparisons, and clinical validation of measurement errors in patients with type 2 neurofibromatosis. PloS one. 2014;9(3):e90300.

16. Huang Wei, Li Xin, Morris Elizabeth A, et al. The magnetic resonance shutter speed discriminates vascular properties of malignant and benign breast tumors in vivo. Proceedings of the National Academy of Sciences. 2008;105(46):17943– 17948.

17. Li Xia, Welch E Brian, Chakravarthy A Bapsi, et al. Statistical comparison of dynamic contrast-enhanced MRI pharmacokinetic models in human breast cancer. Magnetic resonance in medicine. 2012;68(1):261–271.

18. LoCastro Eve, Paudyal Ramesh, Konar Amaresha Shridhar, et al. A Quantitative Multiparametric MRI Analysis Platform for Estimation of Robust Imaging Biomarkers in Clinical Oncology. Tomography. 2023;9(6):2052–2066.

19. Shalom Eve S, Kim Harrison, Der Heijden Rianne A, et al. The ISMRM Open Science Initiative for Perfusion Imaging (OSIPI): Results from the OSIPI–Dynamic Contrast-Enhanced challenge. Magnetic resonance in medicine. 2024;91(5):1803–1821.

20. Woolf David K, Taylor N Jane, Makris Andreas, et al. Arterial input functions in dynamic contrast-enhanced magnetic resonance imaging: which model performs best when assessing breast cancer response?. The British journal of radiology. 2016;89(1063):20150961.

21. Li Xia, Arlinghaus Lori R, Ayers Gregory D, et al. DCEMRI analysis methods for predicting the response of breast cancer to neoadjuvant chemotherapy: Pilot study findings. Magnetic resonance in medicine. 2014;71(4):1592–1602.

22. Ramtohul Toulsie, Tescher Clara, Vaflard Pauline, et al. Prospective evaluation of ultrafast breast MRI for predicting pathologic response after neoadjuvant therapies. Radiology. 2022;305(3):565–574.

23. Thawani Rajat, Gao Lina, Mohinani Ajay, et al. Quantitative DCE-MRI prediction of breast cancer recurrence following neoadjuvant chemotherapy: a preliminary study. BMC Medical Imaging. 2022;22(1):182.

24. Liang Xinhong, Chen Xiaofeng, Yang Zhiqi, et al. Early prediction of pathological complete response to neoadjuvant chemotherapy combining DCE-MRI and apparent diffusion coefficient values in breast Cancer. BMC cancer. 2022;22(1):1250.

25. Tofts Paul S. Modeling tracer kinetics in dynamic GdDTPA MR imaging. Journal of magnetic resonance imaging. 1997;7(1):91–101.

26. Tofts Paul S, Brix Gunnar, Buckley David L, et al. Estimating kinetic parameters from dynamic contrast-enhanced T1-weighted MRI of a diffusable tracer: standardized quantities and symbols. Journal of Magnetic Resonance Imaging: An Official Journal of the International Society for Magnetic Resonance in Medicine. 1999;10(3):223–232.

27. Monge Gaspard. Mémoire sur la théorie des déblais et des remblais. Mem. Math. Phys. Acad. Royale Sci.. 1781;:666– 704.

28. Villani Cédric. Topics in Optimal Transportation. American Mathematical Soc.; 2003.

29. Benamou Jean-David, Brenier Yann. A computational fluid mechanics solution to the Monge-Kantorovich mass transfer problem. Numerische Mathematik. 2000;84(3):375–393.

30. Cuturi Marco. Sinkhorn distances: Lightspeed computation of optimal transport. Advances in neural information processing systems. 2013;26.

31. Benamou Jean-David, Carlier Guillaume, Cuturi Marco, Nenna Luca, Peyré Gabriel. Iterative Bregman projections for regularized transportation problems. SIAM Journal on Scientific Computing. 2015;37(2):A1111–A1138.

32. Chen Yongxin, Georgiou Tryphon T, Pavon Michele. On the relation between optimal transport and Schrödinger bridges: A stochastic control viewpoint. Journal of Optimization Theory and Applications. 2016;169(2):671–691.

33. Léonard Christian. From the Schrödinger problem to the Monge–Kantorovich problem. Journal of Functional Analysis. 2012;262(4):1879–1920.

34. Khamis Abdelwahed, Tsuchida Russell, Tarek Mohamed, Rolland Vivien, Petersson Lars. Scalable Optimal Transport Methods in Machine Learning: A Contemporary Survey. IEEE Transactions on Pattern Analysis and Machine Intelligence. 2024;.

35. Wang Yue, Sun Yongbin, Liu Ziwei, Sarma Sanjay E, Bronstein Michael M, Solomon Justin M. Dynamic graph cnn for learning on point clouds. ACM Transactions on Graphics (tog). 2019;38(5):1–12.

36. Sarlin Paul-Edouard, DeTone Daniel, Malisiewicz Tomasz, Rabinovich Andrew. Superglue: Learning feature matching with graph neural networks. In: :4938–4947; 2020.

37. Balaji Yogesh, Chellappa Rama, Feizi Soheil. Robust optimal transport with applications in generative modeling and domain adaptation. Advances in Neural Information Processing Systems. 2020;33:12934–12944.

38. Moriel Noa, Senel Enes, Friedman Nir, Rajewsky Nikolaus, Karaiskos Nikos, Nitzan Mor. NovoSpaRc: flexible spatial reconstruction of single-cell gene expression with optimal transport. Nature protocols. 2021;16(9):4177–4200.

39. Cang Zixuan, Zhao Yanxiang, Almet Axel A, et al. Screening cell–cell communication in spatial transcriptomics via collective optimal transport. Nature methods. 2023;20(2):218–228.

40. Elkin Rena, Nadeem Saad, Haber Eldad, et al. GlymphVIS: visualizing glymphatic transport pathways using regularized optimal transport. In: :844–852Springer; 2018.

41. Koundal Sunil, others. Optimal Mass Transport with Lagrangian Workflow Reveals Advective and Diffusion Driven Solute Transport in the Glymphatic System. Scientific Reports. 2020;10.

42. Chen Xinan, Tran Anh Phong, Elkin Rena, Benveniste Helene, Tannenbaum Allen R.. Visualizing fluid flows via regularized optimal mass transport with applications to neuroscience. Journal of Scientfic Computing. 2023;97:26.

43. Chen Xinan, Liu Xiaodan, Koundal Sunil, et al. Cerebral amyloid angiopathy is associated with glymphatic transport reduction and time-delayed solute drainage along the neck arteries. Nature aging. 2022;2(3):214–223.

44. Robert Stephanie M., others. The choroid plexus links innate immunity to CSF dysregulation in hydrocephalus. Cell. 2023;186(4):764-785.e21.

45. Ozturk Burhan, Koundal Sunil, Al Bizri Ehab, et al. Continuous positive airway pressure increases CSF flow and glymphatic transport. JCI insight. 2023;8(12).

46. Chen Xinan, Benveniste Helene, Tannenbaum Allen Robert. Unbalanced Regularized Optimal Mass Transport with Applications to Fluid Flows in the Brain. Scientic Reports. 2024;14:1111.

47. Benamou Jean-David. Numerical resolution of an “unbalanced” mass transport problem. ESAIM: Mathematical Modelling and Numerical Analysis. 2003;37(5):851–868.

48. Séjourné Thibault Peyré Gabriel Vialard François-Xavier. Unbalanced optimal transport, from theory to numerics. Handbook of Numerical Analysis. 2023;24:407–471.

49. Chizat Lenaic. Unbalanced optimal transport: Models, numerical methods, applications. PhD thesisUniversité Paris sciences et lettres2017.

50. Feydy Jean, Charlier Benjamin, Vialard François-Xavier, Peyré Gabriel. Optimal transport for diffeomorphic registration. In: :291–299Springer; 2017.

51. Lee John, Bertrand Nicholas P, Rozell Christopher J. Unbalanced optimal transport regularization for imaging problems. IEEE Transactions on Computational Imaging. 2020;6:1219–1232.

52. Schiebinger Geoffrey, Shu Jian, Tabaka Marcin, et al. Optimal-transport analysis of single-cell gene expression identifies developmental trajectories in reprogramming. Cell. 2019;176(4):928–943.

53. Cao Kai, Gong Qiyu, Hong Yiguang, Wan Lin. A unified computational framework for single-cell data integration with optimal transport. Nature Communications. 2022;13(1):7419.

54. Chen Xinan, Huang Wei, Tannenbaum Allen, Deasy Joseph O. Unbalanced Regularized Optimal Mass Transport Theory Applied to Quantify Breast Cancer Tumor Flow Changes on DCE-MRI before and after Neoadjuvant Chemotherapy. In: AAPM; 2023.

55. Chen Xinan, Huang Wei, Tannenbaum Allen R, Deasy Joseph O. Characterizing fluid flows in breast tumor DCEMRI studies using unbalanced regularized optimal mass transport methods. In: SPIE; 2024.

56. Huang Wei, Li Xin, Chen Yiyi, et al. Variations of dynamic contrast-enhanced magnetic resonance imaging in evaluation of breast cancer therapy response: a multicenter data analysis challenge. Translational oncology. 2014;7(1):153–166.

57. Fedorov Andriy, Beichel Reinhard, Kalpathy-Cramer Jayashree, et al. 3D Slicer as an image computing platform for the Quantitative Imaging Network. Magnetic resonance imaging. 2012;30(9):1323–1341.

58. Chizat Lenaic, Peyré Gabriel Schmitzer Bernhard, Vialard Francois-Xavier. An interpolating distance between optimal transport and Fisher-Rao metrics. Foundations of Computational Mathematics. 2016;10:1–44.

59. Buckley David L, Parker Geoffrey JM. Measuring contrast agent concentration in T1-weighted dynamic contrastenhanced MRI. In: Springer 2005 (pp. 69–79).

60. Rathi Yogesh, Olver Peter, Sapiro Guillermo, Tannenbaum Allen. Affine invariant surface evolutions for 3D image segmentation. Image Processing: Algorithms and Systems, Neural Networks, and Machine Learning. 2006;6064:606401.

61. Chen Xinan (Nancy). Unbalanced Regularized Optimal Mass Transport (urOMT). Zenodo. 2023;.

62. Fluckiger Jacob U, Loveless Mary E, Barnes Stephanie L, Lepage Martin, Yankeelov Thomas E. A diffusion-compensated model for the analysis of DCE-MRI data: theory, simulations and experimental results. Physics in Medicine & Biology. 2013;58(6):1983.

63. Sinno Noha, Taylor Edward, Hompland Tord, Milosevic Michael, Jaffray David A, Coolens Catherine. Incorporating cross-voxel exchange for the analysis of dynamic contrast-enhanced imaging data: pre-clinical results. Physics in Medicine & Biology. 2022;67(24):245013.

